# Machine learning enables new insights into clinical significance of and genetic contributions to liver fat accumulation

**DOI:** 10.1101/2020.09.03.20187195

**Authors:** Mary E. Haas, James P. Pirruccello, Samuel N. Friedman, Connor A. Emdin, Veeral H. Ajmera, Tracey G. Simon, Julian R. Homburger, Xiuqing Guo, Matthew Budoff, Kathleen E. Corey, Alicia Y. Zhou, Anthony Philippakis, Patrick T. Ellinor, Rohit Loomba, Puneet Batra, Amit V. Khera

**Affiliations:** Program in Medical and Population Genetics, Broad Institute, Cambridge, MA, 02142; Department of Molecular Biology, Department of Medicine, Massachusetts General Hospital, Boston, MA 02114; Center for Genomic Medicine, Department of Medicine, Massachusetts General Hospital, Boston, MA 02114; Department of Medicine, Harvard Medical School, Boston, MA 02114; Cardiology Division, Department of Medicine, Massachusetts General Hospital, Boston, MA 02114; Data Sciences Platform, Broad Institute, Cambridge, MA, 02142; NAFLD Research Center, Department of Medicine, University of California San Diego, La Jolla, California 92103; Liver Center, Division of Gastroenterology, Department of Medicine, Massachusetts General Hospital, Boston 02114; Color Genomics, Burlingame, CA 94010; The Lundquist Institute for Biomedical Innovation and Department of Pediatrics, Harbor-UCLA Medical Center, Torrance, California 90502

**Author notes:** Corresponding author: Amit V. Khera, MD MSc Center for Genomic Medicine Massachusetts General Hospital 185 Cambridge Street | CPZN 6.256 Boston, MA 02114. Drs. Haas, Pirruccello, and Friedman contributed equally to this manuscript.

## Abstract

Excess accumulation of liver fat – termed hepatic steatosis when fat accounts for > 5.5% of liver content – is a leading risk factor for end-stage liver disease and is strongly associated with important cardiometabolic disorders. Using a truth dataset of 4,511 UK Biobank participants with liver fat previously quantified via abdominal MRI imaging, we developed a machine learning algorithm to quantify liver fat with correlation coefficients of 0.97 and 0.99 in hold-out testing datasets and applied this algorithm to raw imaging data from an additional 32,192 participants. Among all 36,703 individuals with abdominal MRI imaging, median liver fat was 2.2%, with 6,250 (17%) meeting criteria for hepatic steatosis. Although individuals afflicted with hepatic steatosis were more likely to have been diagnosed with conditions such as obesity or diabetes, a prediction model based on clinical data alone without imaging could not reliably estimate liver fat content. To identify genetic drivers of variation in liver fat, we first conducted a common variant association study of 9.8 million variants, confirming three known associations for variants in the *TM6SF2, APOE*, and *PNPLA3* genes and identifying five new variants associated with increased hepatic fat in or near the *MARC1, ADH1B, TRIB1, GPAM* and *MAST3* genes. A polygenic score that integrated information from each of these eight variants was strongly associated with future clinical diagnosis of liver diseases. Next, we performed a rare variant association study in a subset of 11,021 participants with gene sequencing data available, identifying an association between inactivating variants in the *APOB* gene and substantially lower LDL cholesterol, but more than 10-fold increased risk of steatosis. Taken together, these results provide proof of principle for the use of machine learning algorithms on raw imaging data to enable epidemiologic studies and genetic discovery.

## Introduction

Hepatic steatosis – a condition defined by liver fat content > 5.5% – is a leading risk factor for chronic liver disease and is strongly associated with a range of cardiometabolic conditions (Allen et al., 2018; Caussy et al., 2018; Lorbeer et al., 2017; Speliotes et al., 2010). Recent studies have suggested a prevalence of up to 25% across global populations, with rates rapidly increasing in step with the global epidemics of obesity and diabetes (Loomba and Sanyal, 2013; Younossi et al., 2019). Although the condition is frequently undiagnosed within clinical practice, an existing evidence base indicates that avoidance of excessive alcohol intake, weight loss strategies including bariatric surgery, and emerging pharmacologic therapies can each reduce liver fat and prevent progression to more advanced liver disease (Chalasani et al., 2018).

Previous studies related to hepatic steatosis suggest that systematic quantification in large cohorts may prove important in identifying new biologic insights or enabling enhanced clinical care, but suffer from important limitations. First, the traditional approach dichotomizes individuals with hepatic steatosis into ‘nonalcoholic fatty liver disease (NAFLD)’ or alcoholic fatty liver disease according to a largely arbitrary threshold of > 21 drinks per week in men and > 14 drinks per week in women (Chalasani et al., 2018; Sanyal et al., 2011). Second, studies on the prevalence or clinical significance of hepatic steatosis have often been based on non-quantitative ultrasound assessments or physician diagnosis codes, both of which are known to introduce imprecision into downstream analyses (Alexander et al., 2018; Sanyal, 2018; Zhang et al., 2018). Third, common variant association studies (CVAS), which enable systematic comparison of liver fat for individuals stratified by many common variants scattered across the genome, have been hampered by the time-consuming nature of quantifying liver fat from abdominal CT or MRI images and thus have analyzed only up to 14,440 individuals (Speliotes et al., 2011; Kozlitina et al., 2014; Parisinos et al., 2020). By comparison, a recent CVAS of body-mass index – a quantitative trait more easily measured in clinical practice – analyzed up to 681,275 individuals (Yengo et al., 2018).

Based on these prior results, three key areas of uncertainty remain. First, the extent to which a machine learning algorithm can be trained to accurately quantify liver fat in a large group of individuals warrants additional study. Second, the association of clinical risk factors with rates of hepatic steatosis, as well as the ability to predict liver fat content without direct imaging, have not been fully characterized. Third, whether an expanded set of individuals with precise liver fat quantification can enable new genetic discoveries using CVAS or rare variant association studies (RVAS) is largely unknown.

Here we address each of these areas of uncertainty by studying 36,703 middle-aged UK Biobank participants with extensive linked imaging, genetic, and clinical data, the largest such study to date (Figure 1). We develop a machine learning algorithm that precisely quantifies liver fat using raw abdominal MRI images, achieving correlation coefficients of 0.97 and 0.99 in holdout testing datasets. Using this data, we quantify significantly increased rates of hepatic steatosis among key participant subgroups, such as those with obesity or afflicted by diabetes. Genetic analysis identified 8 common DNA variants at genome-wide levels of statistical prevalence or clinical significance of hepatic steatosis have often been based on non-significance, 5 of which were previously unreported, and rare variants in the gene encoding apolipoprotein B (*APOB*) that associate with significantly increased liver fat and rates of steatosis.

**Figure 1.**
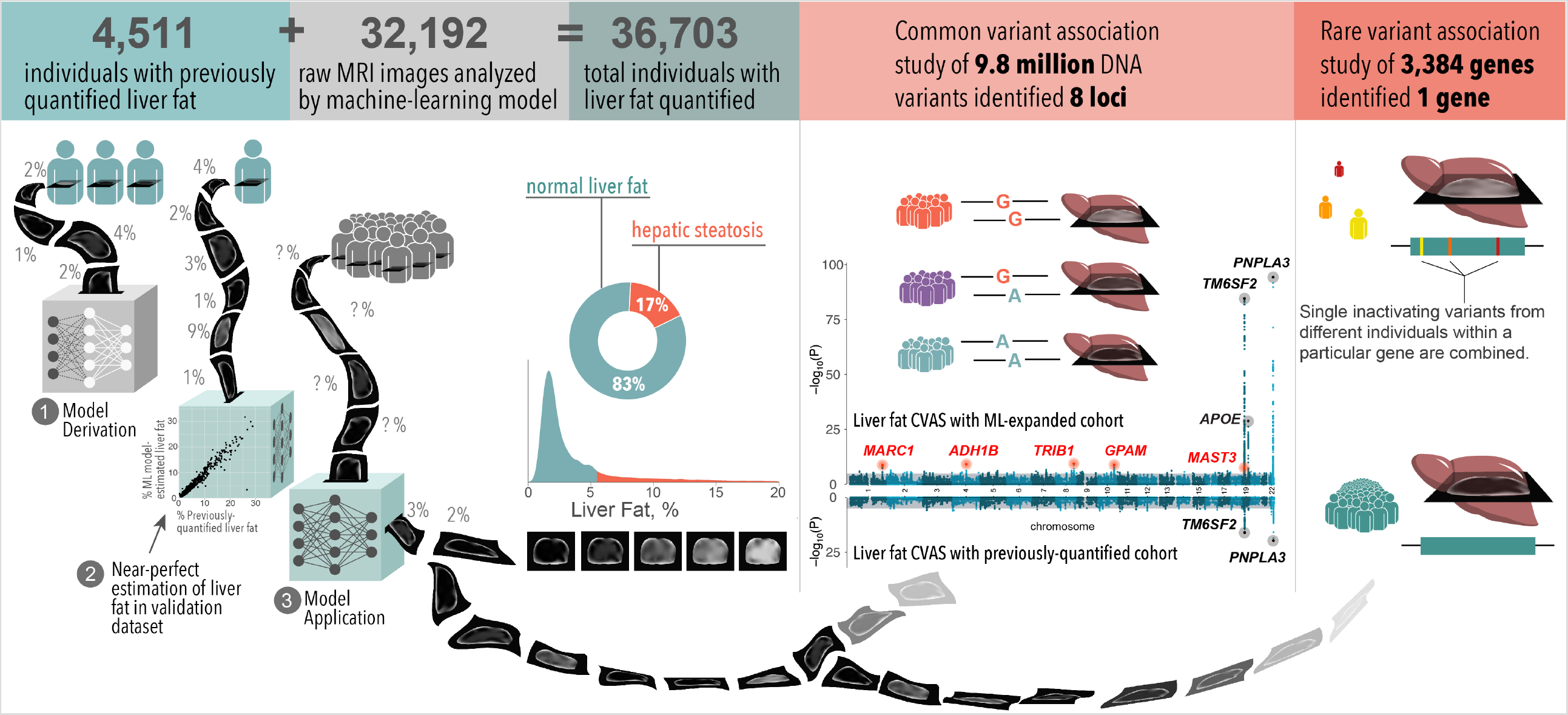
Machine learning enables liver fat quantification, clinical, and genetic analyses. Starting with 4,511 individuals with previously-quantified liver fat, a machine learning model was created to estimate liver fat in an additional 32,192 individuals. Of the 36,703 individuals with resulting liver fat quantification, 17% met criteria for hepatic steatosis. 1.6% had liver fat > 20% (not shown in density plot). A common variant association study (CVAS) identified eight loci, of which five are newly-identified relative to previous CVAS (top Manhattan plot). By contrast, none of these newly-identified variants were identified in a CVAS of those with liver fat previously quantified (bottom Manhattan plot). A rare variant association study identified one gene, *APOB*, in which inactivating variants were significantly associated with liver fat and rates of steatosis.

## Results

### Machine learning model enables near-perfect quantification of hepatic fat based on raw abdominal MRI imaging

To study liver fat in 36,073 UK Biobank participants, we first developed a machine learning algorithm that allowed for precise quantification based on raw abdominal MRI imaging data. We downloaded available imaging data and processed them within a cloud-based computational environment, leveraging a truth dataset of 4,511 participants with liver fat previously quantified by Perspectum Diagnostics (Wilman et al., 2017). Using a two-stage method with deep convolutional neural networks (see methods for details), we trained a machine learning algorithm to quantify liver fat that achieved near-perfect quantification (correlation coefficients 0.97 and 0.99 in hold-out testing datasets in the two stages, Supp. Figure 1). Having trained and validated this algorithm, we next applied this model to quantify liver fat in the remaining 32,192 UK Biobank participants with raw images only.

### Liver fat is strongly associated with cardiometabolic diseases, but cannot be reliably predicted within clinical practice without direct imaging

Across all 36,703 participants studied, median liver fat was 2.2% and 6,250 (17.0%) had liver fat > 5.5% consistent with hepatic steatosis. Mean age at time of imaging was 64 years (range 45 to 82) and 52% were female (Supp. Table 1). Liver fat was significantly increased in male participants versus female (median 2.7 versus 2.0%), those who reported alcohol consumption in excess of current clinical guidelines (median 2.6 versus 2.2%), and those with diagnosed diabetes (median 4.9 versus 2.2%); p-values for each comparison < 0.0001. As expected, median liver fat was significantly higher among 93 individuals with a diagnosis of nonalcoholic fatty liver disease (NAFLD) in the electronic health record as compared to the remainder of the population, median 8.6 versus 2.2% respectively (Figure 2). Consistent with this observation, 56 of 93 (60%) of those diagnosed with NAFLD met imaging-based criteria for hepatic steatosis versus 6,194 of 36,610 (17%) in the remainder of the population, corresponding to an adjusted odds ratio of 7.67 (95%CI 5.03 to 11.70; p-value < 0.0001).

**Figure 2.**
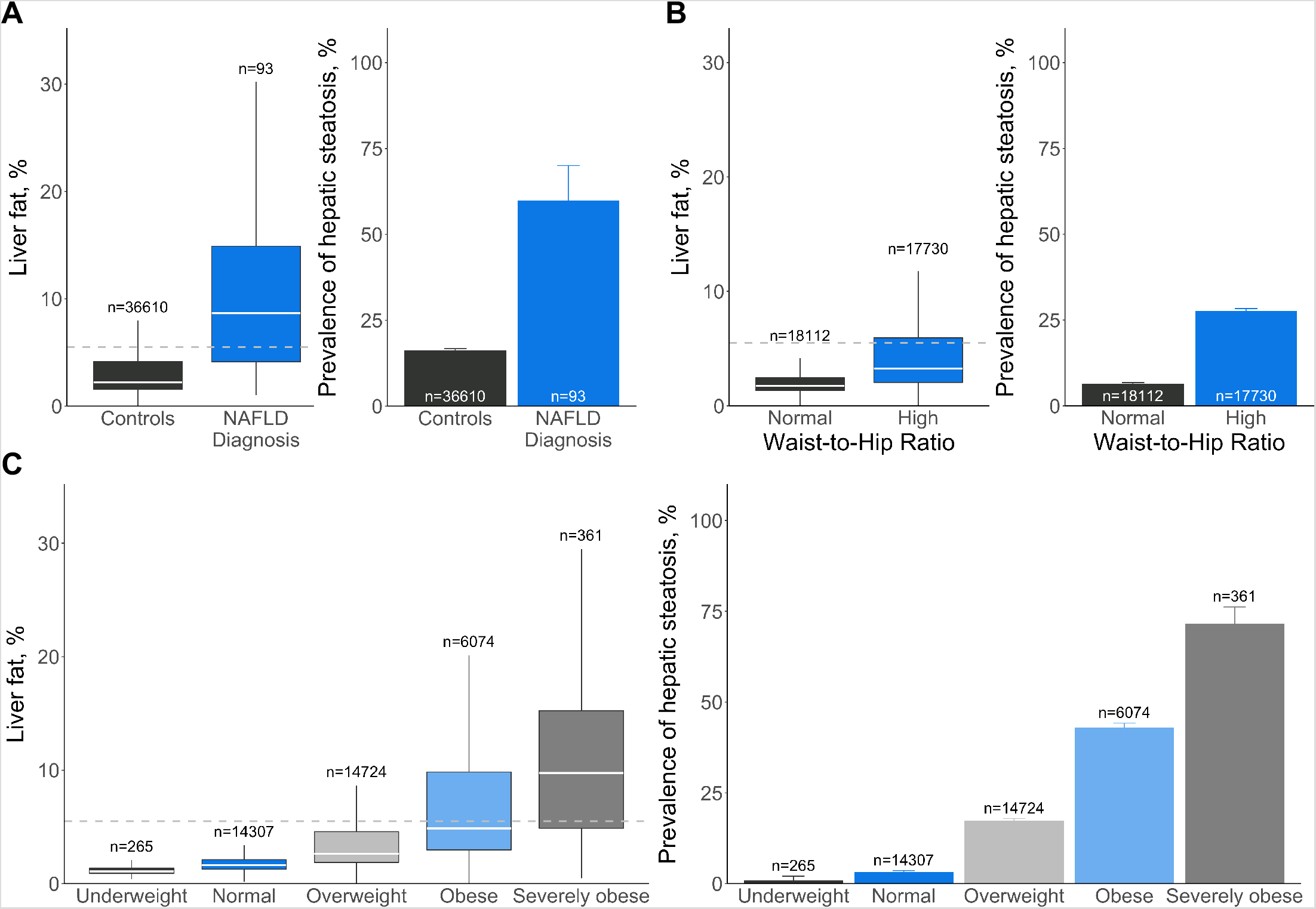
Associations of clinical parameters with liver fat percentage and hepatic steatosis identified by imaging in 36,703 individuals. The distribution of liver fat and prevalence of hepatic steatosis in the presence of A) NAFLD physician diagnosis, B) high waist-to-hip ratio and C) obesity was assessed. Hepatic steatosis was defined as liver fat > 5.5%. High waist-to-hip ratio was defined as greater than 0.9 if male and greater than 0.85 if female at time of imaging. Weight categories were defined using body-mass-index (BMI) at time of imaging: underweight, BMI< 18.5 kg/m^2^; normal, 18.5≤BMI< 25 kg/m^2^; overweight, 25≤BMI< 30 kg/m^2^; obese, 30≤BMI< 40 kg/m^2^; severely obese, BMI≥40 kg/m^2^.

By stratifying individuals according to presence of hepatic steatosis, we observe significant enrichment of cardiometabolic risk factors in those with high liver fat (Table 1). For example, 13.8% of those with steatosis had been diagnosed with diabetes as compared to 3.6% of those in the remainder (adjusted odds ratio 4.21; 95%CI 3.83 to 4.63; p-value < 0.0001), and 45.1% of those with steatosis had been diagnosed with hypertension as compared to 27.1% of those in the remainder (adjusted odds ratio 2.24; 95%CI 2.11 to 2.37; p-value < 0.0001). With respect to biomarkers, circulating triglycerides, liver-associated aminotransferases and glycemic indices were each significantly increased in those with evidence of steatosis.

**Table 1:** Baseline characteristics of UK Biobank participants with quantified liver fat stratified by presence of hepatic steatosis

|  | **Steatosis**<br>**absent**<br>**(N=30,453)** | **Steatosis**<br>**present**<br>**(N=6,250)** | **Unadjusted**<br>**P value** |
| --- | --- | --- | --- |
| Female | 16540 (54.3%) | 2509 (40.1%) | $1 \times 10^{-92}$ |
| Age at enrollment, years | 54.9 (7.51) | 54.7 (7.23) | 0.004 |
| Age at imaging, years | 64.3 (7.62) | 63.8 (7.23) | $9 \times 10^{-7}$ |
| Race |  |  |  |
| White | 29527 (97.0%) | 6045 (96.7%) | 0.355 |
| Black | 185 (0.6%) | 29 (0.5%) | 0.175 |
| South Asian | 239 (0.8%) | 74 (1.2%) | 0.002 |
| Other Asian | 138 (0.5%) | 27 (0.4%) | 0.821 |
| Multiple or other | 280 (0.9%) | 56 (0.9%) | 0.861 |
| Coronary artery disease | 1030 (3.4%) | 253 (4.0%) | 0.009 |
| Diabetes | 1094 (3.6%) | 862 (13.8%) | $2 \times 10^{-234}$ |
| Obese or severely obese | 3964 (13.0%) | 2531 (40.5%) | $< 1 \times 10^{-300}$ |
| Hypertension | 8264 (27.1%) | 2821 (45.1%) | $2 \times 10^{-175}$ |
| Excessive alcohol intake | 1559 (5.1%) | 456 (7.3%) | $6 \times 10^{-12}$ |
| Medications |  |  |  |
| Blood pressure medications | 3555 (11.7%) | 1385 (22.2%) | $2 \times 10^{-108}$ |
| Lipid-lowering therapy | 4287 (14.1%) | 1265 (20.2%) | $3 \times 10^{-35}$ |
| Anthropometric data |  |  |  |
| Weight, kg | 74.7 (13.8) | 86.6 (15.3) | $< 1 \times 10^{-300}$ |
| Waist-to-hip ratio | 0.848 (0.0850) | 0.914 (0.0767) | $< 1 \times 10^{-300}$ |
| Body mass index, kg/m<sup>2</sup> | 25.9 (3.84) | 29.7 (4.41) | $< 1 \times 10^{-300}$ |
| Body fat, % | 29.4 (8.09) | 32.6 (8.10) | $3 \times 10^{-136}$ |
| Systolic blood pressure | 134 (17.6) | 139 (17.0) | $7 \times 10^{-108}$ |
| Weekly drinks, American standard | 5.38 (6.05) | 5.93 (7.75) | 0.021 |
| Liver-associated biomarkers |  |  |  |
| Alanine aminotransferase (IU/L) | 21.3 (12.0) | 31.4 (18.9) | $< 1 \times 10^{-300}$ |
| Aspartate aminotransferase (IU/L) | 25.2 (9.89) | 28.7 (12.9) | $9 \times 10^{-174}$ |
| Gamma glutamyltransferase (IU/L) | 31.3 (31.3) | 45.6 (42.6) | $< 1 \times 10^{-300}$ |
| Lipid biomarkers |  |  |  |
| Total cholesterol, mg/dl | 221 (41.4) | 222 (44.4) | 0.184 |
| LDL cholesterol, mg/dl | 138 (31.7) | 142 (33.5) | $9 \times 10^{-21}$ |
| HDL cholesterol, mg/dL | 58.5 (14.6) | 49.5 (11.6) | $< 1 \times 10^{-300}$ |
| Triglycerides, mg/dL | 116 [84-165] | 171 [122-240] | $< 1 \times 10^{-300}$ |
| Glycemic biomarkers |  |  |  |
| Glycated hemoglobin, % | 5.33 (0.439) | 5.50 (0.601) | $1 \times 10^{-128}$ |
| Glucose, mg/dL | 89.2 (16.0) | 93.4 (23.1) | $6 \times 10^{-45}$ |
| Liver fat at imaging, % | 1.96 [1.47-2.85] | 9.86 [7.13-14.24] | $< 1 \times 10^{-300}$ |

Values correspond to number (%), mean (standard deviation), or median [interquartile range]. P-values correspond to chi-squared test or Wilcoxon rank sum for categorical and continuous variables, respectively.

Despite the correlation of liver fat with cardiometabolic risk factors, practicing clinicians would not be able to reliably estimate liver fat without direct imaging assessment. As an example, the Pearson correlation coefficient between body mass index – the most commonly used surrogate for adiposity – and liver fat was 0.48, but a broad range of observed values were observed across categorizations used in clinical practice (Figure 2). For those with severe obesity (body-mass index ≥ 40 kg/m^2^), median liver fat was 9.8% and 107 of 361 (70%) met clinical criteria for steatosis, but measured liver fat ranged widely from 0.5 to 31.5%. Even for those with normal weight in whom median liver fat was 1.6%, 470 of 14,307 (3%) nonetheless had imaging evidence of hepatic steatosis. Similarly, only 4,854 of 17,730 (27%) with a high waist-to-hip ratio, a measure of central adiposity, had liver fat consistent with hepatic steatosis. To further assess the ability to predict liver fat using variables available in clinical practice, we built a beta regression model using 25 variables, including measures of adiposity and blood lipids and biomarkers of liver injury and assessed its predictive accuracy in held out participants, noting a Pearson correlation coefficient of only 0.45 (Supp. Figure 2).

### A common variant association study identifies 8 genetic variants – including 5 newly-identified – associated with increased liver fat

We first confirmed prior studies noting a significant inherited component to liver fat (Speliotes et al., 2011; Palmer et al., 2013; Loomba et al., 2015), estimating that up to 33% of the observed variance is explained by measured genetic variants when considered in aggregate using the BOLT-REML method (Loh et al., 2015a2015a). To identify the specific variants most strongly contributing to this heritability, we performed a common variant association study (CVAS), assessing the relationship of each of 9.8 million common (minor allele frequency ≥ 1%) DNA variants and liver fat percentage. Given that 97% of individuals with liver fat quantified were of European ancestry (Supp. Table 1) and the potential for small numbers of individuals of distinct ancestries to introduce confounding by population stratification, we restricted our CVAS to 32,976 individuals of European ancestries using a combination of self-reported ethnicity and genetic principal component analysis (Bycroft et al., 2018, see Methods). Moreover, because standard CVAS algorithms include an assumption that the residuals of the phenotype are normally distributed, we applied an inverse-normal transformation to liver fat residuals prior to CVAS resulting in a Gaussian distribution with mean of zero and standard deviation of one.

The CVAS identified 8 loci in which common genetic variants were associated with increased liver fat, including 5 not previously identified at genome-wide levels of statistical significance (Figure 3, Table 2). We confirm and extend prior observations for missense variants in the *TM6SF2, PNPLA3*, and *APOE* genes. Within units of the inverse-normal transformed liver fat variable, the impact of these variants ranged from +0.12 to +0.29 standard deviations, corresponding to impact on raw liver fat percentage ranging from 0.51 to 1.37% and odds ratio of hepatic steatosis per allele of 1.40 to 1.90.

**Figure 3.**
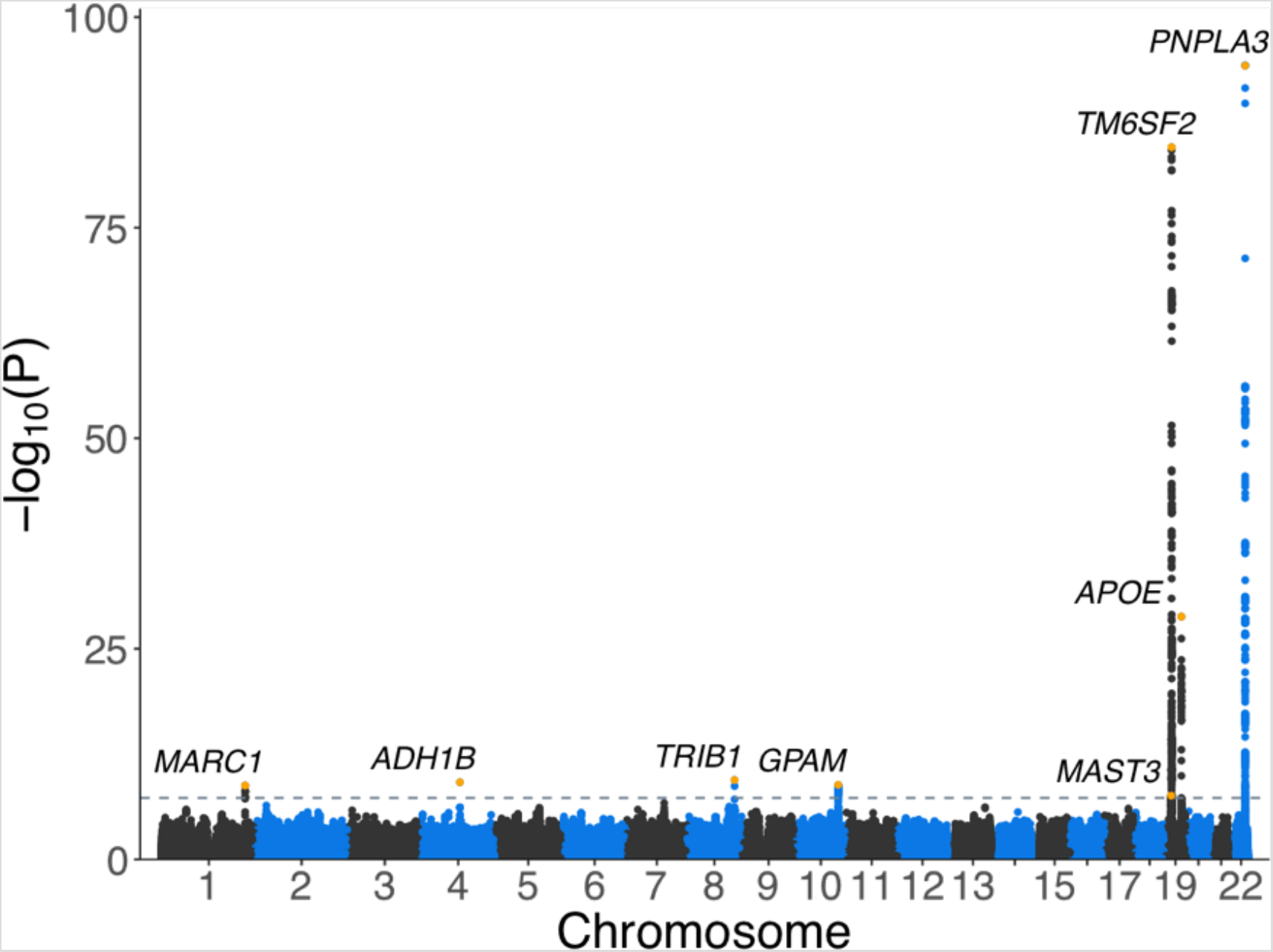
Genome-wide association study of liver fat in UK Biobank identifies 8 loci. The lead variant at each of 8 genome-wide significant loci are indicated by orange points. Gray line indicates genome-wide significance threshold (p< 5×10^−8^).

**Table 2:** 8 common DNA variants associated with liver fat indices

| Lead Variant | Chr. | Position (hg19) | Nearest Gene | Consequence | Effect Allele | Other Allele | Effect Freq. | Effect on liver fat (SD) (95%CI)<br>p value | Effect on % liver fat (95%CI)<br>p value | Effect on hepatic steatosis (OR) (95%CI)<br>p value) |
| --- | --- | --- | --- | --- | --- | --- | --- | --- | --- | --- |
| **Newly-identified variants** |  |  |  |  |  |  |  |  |  |  |
| rs2642438 | 1 | 220970028 | *MARC1* | Missense (p.T165A) | G | A | 0.70 | 0.05<br>(0.04-0.07)<br>p=2x10<sup>-9</sup> | 0.22<br>(0.14-0.29)<br>p=3x10<sup>-9</sup> | 1.17<br>(1.11-1.22)<br>p=6x10<sup>-11</sup> |
| rs1229984 | 4 | 100239319 | *ADH1B* | Missense (p.H48R) | C | T | 0.98 | 0.16<br>(0.11-0.21)<br>p=7x10<sup>-10</sup> | 0.51<br>(0.29-0.72)<br>p=3x10<sup>-6</sup> | 1.37<br>(1.18-1.59)<br>p=3x10<sup>-5</sup> |
| rs112875651 | 8 | 126506694 | *TRIB1* | Intergenic | G | A | 0.61 | 0.05<br>(0.03-0.07)<br>p=4x10<sup>-10</sup> | 0.19<br>(0.13-0.26)<br>p=2x10<sup>-8</sup> | 1.10<br>(1.06-1.15)<br>p=9x10<sup>-6</sup> |
| rs2250802 | 10 | 113921354 | *GPAM* | Intronic | G | A | 0.27 | 0.05<br>(0.04-0.07)<br>p=1x10<sup>-9</sup> | 0.24<br>(0.17-0.31)<br>p=1x10<sup>-10</sup> | 1.13<br>(1.08-1.18)<br>p=1x10<sup>-7</sup> |
| rs56252442 | 19 | 18229208 | *MAST3* | Intronic | T | G | 0.25 | 0.05<br>(0.03-0.07)<br>p=3x10<sup>-8</sup> | 0.18<br>(0.1-0.25)<br>p=3x10<sup>-6</sup> | 1.09<br>(1.04-1.14)<br>p=3x10<sup>-4</sup> |
| **Previously-identified variants** |  |  |  |  |  |  |  |  |  |  |
| rs58542926 | 19 | 19379549 | *TM6SF2* | Missense (p.E167K) | T | C | 0.07 | 0.29<br>(0.26-0.32)<br>p=3x10<sup>-85</sup> | 1.37<br>(1.25-1.49)<br>p=1x10<sup>-104</sup> | 1.90<br>(1.78-2.04)<br>p=1x10<sup>-75</sup> |
| rs429358 | 19 | 45411941 | *APOE* | Missense (p.R130C) | T | C | 0.85 | 0.12<br>(0.10-0.14)<br>p=2x10<sup>-29</sup> | 0.51<br>(0.42-0.60)<br>p=2x10<sup>-28</sup> | 1.40<br>(1.32-1.49)<br>p=2x10<sup>-26</sup> |
| rs738409<sup>a</sup> | 22 | 44324727 | *PNPLA3* | Missense (p.I148M) | G | C | 0.21 | 0.19<br>(0.18-0.21)<br>p=6x10<sup>-95</sup> | 0.88<br>(0.81-0.96)<br>p=1x10<sup>-106</sup> | 1.59<br>(1.52-1.66)<br>p=7x10<sup>-83</sup> |

<sup>a</sup>rs738409, the known causal variant in the *PNPLA3* gene region (Romeo et al., 2008) is in near-perfect disequilibrium (inherited together, R<sup>2</sup>=0.999) with the lead variant in our study, rs738408. Chr., chromosome; Freq., frequency; SD, standard deviation; OR, odds ratio.

Genetic variants in the five newly-identified loci included: the p.T165A missense variant in the gene encoding Mitochondrial Amidoxime Reducing Component 1 (*MARC1*), which we and others recently identified as associated with end-stage liver disease (Emdin et al., 2020; Innes et al., 2020; Luukkonen et al., 2020); the p.H48R missense variant in the gene encoding Alcohol Dehydrogenase 1B (Class I), Beta Polypeptide (*ADH1B*), which plays a key role in ethanol oxidation to acetaldehyde, is thought to contribute to aversion to alcohol intake and was previously associated with alcohol consumption (Bierut et al., 2012; Walters et al., 2018); an intergenic variant near the gene encoding Tribbles-Like Protein 1 (*TRIB1*), which is known to associate with circulating triglycerides and play a role in hepatic lipogenesis (Kathiresan et al., 2008; Burkhardt et al., 2010; Bauer et al., 2015); an intronic variant in the gene encoding glycerol-3-phosphate acyltransferase, mitochondrial (*GPAM*), previously linked to liver triglyceride content in murine overexpression and knockout experiments (Hammond et al., 2002; Lindén et al., 2006); and an intronic variant in the gene encoding microtubule associated serine/threonine kinase 3 (*MAST3*), previously linked to inflammatory bowel disease (Labbé et al., 2008) but with as-yet unknown role in liver fat metabolism. Among these five newly-identified variants, impact on liver fat percentage ranged from 0.18 to 0.51% and odds ratio for hepatic steatosis per allele ranged from 1.09 to 1.37 (Table 2).

The expansion of individuals with liver fat quantification from 4,038 to 32,976 individuals using machine learning played a key role in enabling the CVAS. Taking the most strongly associated variant – the p.I148M missense variant in *PNPLA3* – as an example, the p-value for association decreased from 2×10^−20^ when performing a CVAS in only 4,038 individuals to 5.6×10^−95^ when using 32,976 participants. Moreover, although each of the five newly-identified variants had directionally-consistent evidence of association in a CVAS of 4,038 individuals (pvalues ranging from 0.16 to 3.6×10^−4^) none met the standard threshold for statistical significance of p = 5×10^−8^.

Beyond variants meeting genome-wide levels of statistical significance, we next sought to replicate additional variants previously reported to affect liver fat or risk of NAFLD (Supp. Table 3). A missense variant in the gene encoding the glucokinase receptor (*GCKR*) (Speliotes et al., 2011; Palmer et al., 2013; Parisinos et al., 2020) was strongly associated with liver fat with subthreshold level of statistical significance (p = 4.1×10^−7^), as was a variant near the gene encoding Membrane Bound O-Acyltransferase Domain Containing 7 (*MBOAT7*; p = 8.8×10^−6^; Buch et al., 2015; Mancina et al., 2016). Consistent with prior reports suggesting that an inactivating variant in the gene encoding hydroxysteroid 17-beta dehydrogenase 13 (*HSD17B13*) relates more strongly to progression from steatosis to more advanced forms of liver disease, we did not observe an association with liver fat in our study population (p = 0.40, Abul-Husn et al., 2018; Ma et al., 2019; Gellert-Kristensen et al., 2020).

Given a known important role of alcohol intake on liver fat, we next performed two sensitivity analyses: first, we repeated the CVAS after exclusion of 3,013 individuals who reported alcohol consumption in excess of current clinical recommendations or who reported having stopped drinking alcohol; and second, we repeated the CVAS including self-reported number of alcoholic drinks consumed per week as a covariate. In both cases, results for the 8 variants identified were highly consistent, suggesting that these variants have a largely consistent impact on liver fat independent of alcohol consumption (Supp. Table 4).

To replicate the CVAS associations in additional independent cohorts, we analyzed liver fat as assessed by an alternate imaging modality (computed tomography) in 3,284 participants of the Framingham Heart Study and 4,195 participants of the Multi-Ethnic Study of Atherosclerosis (MESA) study. In the Framingham Heart Study, the average age at time of imaging was 52 and 48% were female; in MESA, the average age was 61 and 51% were female. Although the computed tomography measures of hepatic fat based on liver attenuation cannot be directly converted to liver fat percentage, all 8 variants’ associations were directionally consistent and 5 were nominally significant (p< 0.05, Supp. Table 5).

Beyond association with liver fat indices, we assessed for additional validation of the variants identified by CVAS using liver biomarkers and clinical diagnoses entered into the medical record. In UK Biobank, we analyzed up to 362,910 UK Biobank participants, excluding those included in the abdominal MRI substudy. We first determined associations with the blood biomarkers alanine aminotransferase (ALT) and aspartate aminotransferase (AST). All eight variants were robustly associated with increased ALT, and 7 of the 8 variants were associated with increased AST at nominal levels of statistical significance (p < 0.05, Supp. Table 6). We next examined association of the CVAS variants with a recorded clinical diagnosis of NAFLD or nonalcoholic steatohepatitis (NASH), a more advanced form of fatty liver disease that additionally includes significant liver inflammation, in both the UK Biobank and the Mass General Brigham Biobank, a hospital-based biorepository (Karlson et al., 2016). 2,225 of 362,910 participants in the UK Biobank, and 4,129 of 30,573 participants of the Mass General Brigham Biobank had been diagnosed with NAFLD or NASH. In a meta-analysis of these two studies, 7 of the 8 variants were strongly associated with increased risk, with odds ratio ranging from 1.08 to 1.43 (p < 0.004 for each; Supp. Table 7). The remaining variant, rs56252442 near *MAST3*, was directionally consistent but did not achieve statistical significance (odds ratio 1.02; 95% CI 0.98 to 1.07; p = 0.32), and we thus consider this association a preliminary result warranting further replication.

### A polygenic score comprised of 8 common genetic variants is strongly associated with chronic liver diseases

Recognizing that each of the 8 common variants have modest individual impact on liver fat percentage or risk of steatosis, we next integrated information from each of them into a weighted polygenic score. Within the discovery study population of 32,976 UK Biobank individuals, this polygenic score explained 3.5% of the observed variance. To determine the relationship of this polygenic score to chronic liver diseases, we calculated it in 362,096 UK Biobank participants who were *not* included in the liver fat imaging substudy and had not been diagnosed with liver disease at time of enrollment. The polygenic score was strongly associated with a new diagnosis code of NAFLD entered into the medical record during follow-up, with a hazard ratio per standard deviation score increment (HR/SD) of 1.33 (95%CI 1.27 to 1.39, Figure 4). Individuals who carried a diagnosis of NAFLD had a median polygenic score in the 61st percentile of the distribution as compared to the 50th percentile for those in the remainder of the population. Beyond NAFLD, the polygenic score was additionally associated with an increased risk of more advanced forms of liver disease: nonalcoholic steatohepatitis (HR/SD 1.66), cirrhosis (HR/SD 1.40), and hepatocellular carcinoma (HR/SD 1.69). Based on prior observations of an association between liver disease risk-increasing alleles of variants in the *PNPLA3* and *TM6SF2* genes and decreased cholesterol (Liu et al., 2017), we determined the relationship of the polygenic score to circulating LDL cholesterol. Each SD increment in the score was associated with a 1.9 mg/dl (95%CI 1.8 to 2.0; p = 1.5×10^−244^) decrease in measured LDL cholesterol concentrations, illustrating a tradeoff rooted in rates of hepatic cholesterol secretion with potentially important implications for drug development.

**Figure 4:**
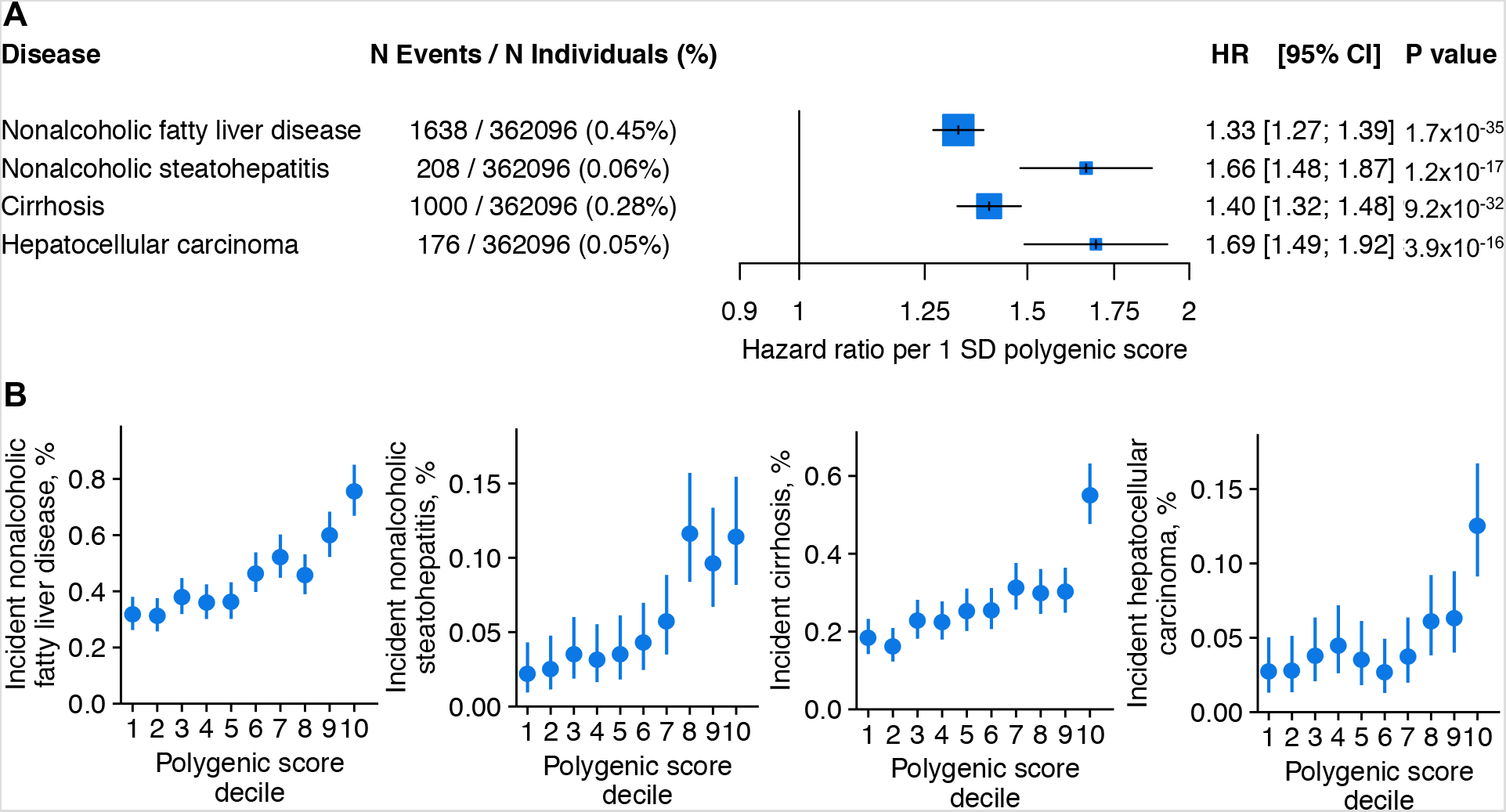
Association between a polygenic score comprised of 8 common DNA variants and risk of liver diseases. The 8-variant polygenic score was used to predict new liver-related disease diagnoses in 362,096 UK Biobank participants who were not included in the discovery CVAS of individuals with imaging data. A) Hazard ratios (HR) of incident disease per 1 standard deviation (SD) increase in the polygenic score; error bars represent 95% confidence intervals (CI). B) Rates of incident disease in each decile of the polygenic score; error bars represent 95% binomial distribution confidence intervals.

### Rare variant association study identifies inactivating variants in *APOB* and *MTTP* that increase liver fat

For the subset of 11,021 UK Biobank participants with both liver fat quantified and gene sequencing available, we next investigated whether rare inactivating DNA variants might similarly impact liver fat or risk of steatosis. Because such inactivating mutations do not occur with adequate frequency to detect individual variant-phenotype relationships, we performed a ‘collapsing burden’ rare variant association study (RVAS). In this approach, the observed liver fat for carriers of any inactivating variant for a given gene is compared to individuals without such an inactivating variant. This analysis was restricted to 3,384 genes with at least 10 carriers of inactivating mutations observed, resulting in a Bonferroni-corrected p-value of statistical significance of 1.48×10^−5^ (p = 0.05/3,384).

Inactivating variants in the gene encoding apolipoprotein B (*APOB*) – known to play a key role in lipid homeostasis – were associated with substantially increased liver fat. Using units of inverse-normal transformed hepatic fat, values were 1.58 standard deviations higher (95%CI 0.91 to 2.24; p = 3.25 × 10^−6^) among 11 carriers of inactivating mutations versus 11,010 individuals without such a variant. This corresponded to a median liver fat of 9.7% versus 2.1% for carriers and noncarriers respectively, and an odds ratio for hepatic steatosis among carriers of 10.8 (95%CI 2.8 to 41.4; p = 5.3×10^−4^) (Figure 5).

**Figure 5.**
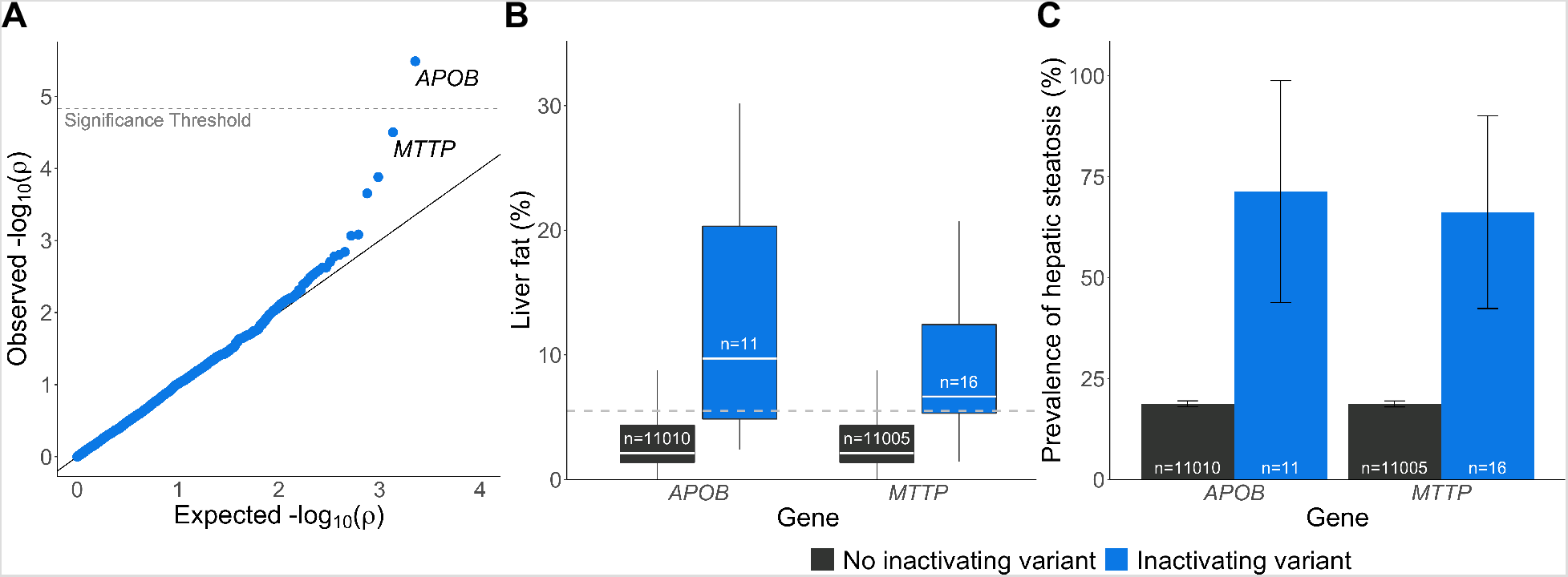
Rare variant association study of liver fat in 11,120 individuals. A) QQ plot with significance threshold indicated (0.05/3384 genes tested = 1.48×10^−5^). B) Liver fat distribution in carriers and noncarriers of inactivating variants in the *APOB* or *MTTP* genes. C) Prevalence of hepatic steatosis (liver fat > 5.5%) in carriers and noncarriers of inactivating variants in *APOB* or *MTTP*.

Significant prior human genetic and pharmacologic data implicate APOB in hepatic fat accumulation. Individuals with two inactivating mutations in *APOB* (‘human knockouts’) suffer from the Mendelian condition familial hypobetalipoproteinemia, characterized by near absent levels of circulating apolipoprotein B and LDL cholesterol but increased rates of hepatic steatosis (Lee and Hegele, 2013). Similarly, pharmacologic knockdown of the *APOB* gene using the antisense oligonucleotide mipomersen is approved for the treatment of severe hypercholesterolemia, but is infrequently used within clinical practice owing to high rates of hepatic steatosis observed in clinical trials.

To further characterize the phenotypic consequences of inactivating variants in *APOB*, we analyzed an expanded set of 42,260 UK Biobank participants with gene sequencing data available (regardless of availability of abdominal MRI imaging data). Of these 42,260 individuals, 64 (0.15%) harbored an inactivating variant in *APOB*. Biomarkers of liver injury were increased in these individuals: 31% higher alanine aminotransferase and 14% higher aspartate aminotransferase, p-values 1.7×10^−5^ and 0.006 respectively (Supp. Table 8). In contrast to higher values of aminotransferases, carriers of inactivating *APOB* variants had markedly lower levels of circulating lipoproteins: 40% lower apolipoprotein B, 44% lower LDL cholesterol, and B C 40% lower triglycerides (p < 0.0001 for each). This reduction in circulating lipids was associated with a 75% reduction in risk of coronary artery disease, which although not reaching statistical significance (p = 0.15), is consistent with our recent report of a 72% lower risk of coronary artery disease (p = 0.002) in a much larger dataset (Peloso et al., 2019).

Across all 3,348 genes, inactivating variants in the gene encoding microsomal triglyceride transfer protein (*MTTP*) demonstrated the second strongest strength of association. Using units of inverse-normal transformed hepatic fat, values were 1.17 standard deviations higher (95%CI 0.62 to 1.72; p = 3.14 × 10^−5^) among 16 carriers of inactivating mutations versus 11,005 individuals without such a variant. This corresponded to a median liver fat percentage of 6.7% versus 2.1% for carriers and noncarriers respectively, and an odds ratio for hepatic steatosis of 8.5 (95%CI 2.9 to 24.8; p = 8.5×10^−5^) for carriers (Figure 5).

Although the association of inactivating variants in *MTTP* with liver fat did not reach our prespecified levels of statistical significance, this observation is highly consistent with known biology. MTTP plays a central role in secretion of apolipoprotein B-containing lipoproteins from the liver. Individuals with two inactivating *MTTP* variants suffer from the Mendelian disorder abetalipoproteinemia, a condition characterized by absence of circulating apolipoprotein B and increased rates of hepatic steatosis (Berriot-Varoqueaux et al., 2000; Sharp et al., 1993). As was noted for APOB inhibition, a pharmacologic inhibitor of MTTP activity is approved for treatment of severe hypercholesterolemia, but clinical use is limited by increased hepatic fat observed after administration (Cuchel et al., 2007). Consistent with prior data suggesting that inactivating *MTTP* variants impact circulating lipids only when both copies are affected via a recessive inheritance pattern (Lee and Hegele, 2013), the individuals we identified with a single inactivating variant did not have significantly reduced lipid concentrations (Supp. Table 8).

## Discussion

Our analysis describing quantification of liver fat in 36,703 middle-aged individuals using a machine learning algorithm trained on a small subset with previously quantified values has several implications for both biologic discovery and clinical medicine.

First, the near-perfect estimation of liver fat enabled by a high-throughput machine learning algorithm confirms and extends prior such efforts and is likely to be broadly generalizable across a diverse spectrum of important phenotypes. Within hold-out testing datasets, our model-based liver fat assessment was highly correlated with liver fat previously quantified by a commercial vendor, with correlation coefficients of 0.97 and 0.99. Previous efforts have similarly documented feasibility of using a convolutional neural net framework to automate liver fat quantification using either CT or MRI images obtained in clinical practice (Wang et al., 2019). Beyond liver phenotypes, we recently validated a machine learning model to quantify the diameter of the aorta – the major vessel supplying oxygen-rich blood to the body – using cardiac MRI imaging data, enabling discovery of 93 associated genetic variants (Pirruccello et al., 2020). Taken together, these and other studies (Meyer et al., 2020) suggest that machine learning approaches to rapidly quantify phenotypes of interest in rich imaging datasets are likely to yield important new scientific insights, particularly if extended to complex features derived from dynamic tissues such as a beating heart, or latent phenotypes not currently measured within clinical practice.

Second, we demonstrate that, although correlated with many cardiometabolic traits, liver fat cannot be readily predicted using information readily available in clinical practice. Our large-scale study confirmed significantly increased liver fat in important clinical subgroups such as those with diabetes or those with severe obesity. These observations suggest that future research might validate a clinical prediction tool – potentially including a polygenic score – that identifies a subgroup of individuals in whom screening for hepatic steatosis is warranted, or those with known steatosis who are most likely to progress to cirrhosis (Ajmera et al., 2018). Even if not performed within the context of focused screening, abdominal imaging is very common across a wide range of clinical indications. Application of a machine learning algorithm to alert ordering clinicians of an incidental finding of hepatic steatosis may enable measures that prevent progression to more advanced liver disease (Chalasani et al., 2018). This approach has proven useful in identifying individuals with subclinical atherosclerosis on chest CT imaging, and reporting as an incidental finding is now recommended by current practice guidelines (Hecht et al., 2017; Pakdaman et al., 2017).

Third, a common variant association study enabled by our machine learning algorithm more than doubled the number of significantly associated variants in our study from 3 to 8. None of the five newly-identified variants were identified using the subset of 4,038 individuals with liver fat quantified without machine learning. We note compelling biology underlying most of the observed variants, and provide proof-of-concept that combining several most strongly associated variants into a polygenic score was associated with risk of liver-related diseases. By providing the full set of summary association statistics for our CVAS at the time of publication, we hope to enable additional downstream research.

Fourth, a rare variant association study, despite a relatively small sample size of 11,021 individuals with both liver fat and gene sequencing data available, identified relationships between inactivating variants in *APOB* and *MTTP* with liver fat. These observations recapitulate the results observed in pharmacologic studies focused on APOB or MTTP inhibition as a treatment for hypercholesterolemia: those with inactivating variants in *APOB* had strikingly lower lipid concentrations, but this came at the expense of increased aminotransferase concentrations and a more than 10.8-fold increase in rates of hepatic steatosis. Given that liver biomarker elevations or increased hepatic fat are commonly observed adverse reactions to novel drug candidates – in many cases leading to termination of drug development programs – our approach to using genetics to predict hepatotoxicity may prove valuable. Moreover, our results suggest that a subset of candidate treatment strategies for hepatic steatosis may have adverse effects on increasing circulating lipids. Seen in this light, prioritization of some drug targets, such as MARC1, where genetic studies suggest inhibition will protect against liver disease without adversely increasing cholesterol concentrations or risk of cardiovascular disease (Emdin et al., 2020), may be warranted.

Our results should be interpreted within the context of several potential limitations. First, participants of the UK Biobank imaging study tend to be healthier than the general population and the majority are of European ancestries; additional research is needed to confirm generalizability and transethnic portability. Second, diagnostic codes entered into the electronic health record were used to study the relationship between a clinical diagnosis of NAFLD and liver fat parameters based on imaging; such codes are known to be imprecise within the context of clinical care. Future studies involving manual chart review or using biopsy-confirmed cases of liver disease are warranted. Third, because imaging occurred very recently, we were not able to explore the relationship between liver fat assessment and future risk of cardiometabolic disease endpoints.

In conclusion, we applied a machine learning algorithm to quantify liver fat in 36,703 participants of the UK Biobank, identifying 17% of the population with evidence of hepatic steatosis despite lack of a recorded clinical diagnosis of fatty liver disease, and enabling new genetic discoveries with potential important implications for identifying new mechanistic pathways underlying risk for liver disease.

## Data Availability

Summary statistics for the liver fat CVAS, as well as the machine learning model architectures and learned weights will be available at the Cardiovascular Disease Knowledge Portal (http://broadcvdi.org/home/portalHome) and the ML4CVD modeling framework will be available via GitHub repository at time of publication.

## Acknowledgements

This research has been conducted using the UK Biobank resource, application 7089. Funding support was provided by NIH grants 1K08HG010155 (to A.V.K.) from the National Human Genome Research Institute, 1R01HL092577, R01HL128914, K24HL105780 (to P.T.E), R01HL071739 (to M.B.) from the National Heart, Lung and Blood Institute, 5P42ES010337 (to R.L.) from the National Institute of Environmental Health Sciences, 5UL1TR001442 (to R.L.) from the National Center for Advancing Translational Sciences, R01DK106419, P30DK120515 (to R.L.), K23 DK122104 to (to T.G.S.) from the National Institute of Diabetes and Digestive and Kidney Diseases, CA170674P2 (to R.L.) from the Department of Defense’s Peer Reviewed Cancer Research Program, a Hassenfeld Scholar Award from Massachusetts General Hospital (to A.V.K.), a Merkin Institute Fellowship from the Broad Institute of MIT and Harvard (to A.V.K.), a John S LaDue Memorial Fellowship (to J.P.P.) a sponsored research agreement from IBM Research (to A.P., A.V.K.), American Association for the Study of Liver Diseases Foundation Clinical and Translational Research Awards (to V.A. and T.G.S.).

MESA and the MESA SHARe projects are conducted and supported by the National Heart, Lung, and Blood Institute (NHLBI) in collaboration with MESA investigators. Support for MESA is provided by contracts 75N92020D00001, HHSN268201500003I, N01-HC-95159, 75N92020D00005, N01-HC-95160, 75N92020D00002, N01-HC-95161, 75N92020D00003, N01-HC-95162, 75N92020D00006, N01-HC-95163, 75N92020D00004, N01-HC-95164, 75N92020D00007, N01-HC-95165, N01-HC-95166, N01-HC-95167, N01-HC-95168, N01-HC-95169, UL1-TR-000040, UL1-TR-001079, UL1-TR-001420, and supported in part by the National Center for Advancing Translational Sciences, CTSI grant UL1TR001881, and the National Institute of Diabetes and Digestive and Kidney Disease Diabetes Research Center (DRC) grant DK063491 to the Southern California Diabetes Endocrinology Research Center. The authors thank the other investigators, the staff, and the participants of the MESA study for their valuable contributions. A full list of participating MESA investigators and institutions can be found at http://www.mesa-nhlbi.org.

## Author Contributions

M.E.H., J.P.P. and S.N.F. conceived of the study. M.E.H., J.P.P., S.N.F. and C.A.E. conducted analyses. M.E.H., J.P.P., S.N.F., and A.V.K. wrote the paper. All other authors contributed to the analysis plan or provided critical revisions.

## Methods

### Study cohorts

#### UK Biobank

The UK Biobank is a prospective cohort study that enrolled 502,617 individuals aged 40–69 years of age from across the United Kingdom (Sudlow et al., 2015). As part of the study protocol, a subset of individuals underwent detailed imaging between years 2014 and 2019 including abdominal MRI (Littlejohns et al., 2020).

We first analyzed 36,703 UK Biobank participants with abdominal MRI imaging. The UK Biobank abdominal imaging protocol was first performed with gradient echo imaging; 4,511 participants had liver fat originally quantified by Perspectum Diagnostics as previously described (Wilman et al., 2017). Beginning in 2018, imaging was switched to the “iterative decomposition of water and fat with echo asymmetry and least-squares estimation” (IDEAL) protocol. A subset of participants underwent both imaging protocols.

We next performed a common variant association study (CVAS) of liver fat on a subset of 32,976 participants. We excluded samples that had no imputed genetic data, a genotyping call rate < .98, a mismatch between submitted and inferred sex, sex chromosome aneuploidy (neither XX nor XY), exclusion from kinship inference, excessive third-degree relatives, or that were outliers in heterozygosity or genotype missingness rates, all of which were previously defined centrally by the UK Biobank (Bycroft et al., 2018). Due to the small percentage of samples of non-European ancestries (Supp. Table 1), to avoid artifacts from population stratification we restricted our GWAS to samples of European ancestries (determined as previously reported (Haas et al., 2018) via self-reported ancestry of British, Irish, or other white and outlier detection using the R package *aberrant*). We did not remove related individuals from this analysis as we used a linear mixed model able to account for cryptic relatedness in common variant association studies (Loh et al., 2015b).

To further validate the common variants associated with liver fat in the CVAS, we studied association of single variants as well as a composite 8-variant polygenic risk score (PRS) with liver disease and/or blood biomarkers alanine aminotransferase (ALT) and aspartate aminotransferase (AST) in 362,910 individuals in the UK Biobank who did not undergo imaging and therefore were not part of the discovery cohort. Sample quality control was performed by excluding samples that had no imputed genetic data, a genotyping call rate < 0.95, a mismatch between submitted and inferred sex, sex chromosome aneuploidy, exclusion from kinship inference, excessive third-degree relatives, or that were outliers in heterozygosity or genotype missingness rates, and restricting to samples of European ancestries. We additionally removed one of each pair of related individuals (2nd degree or closer, KING coefficient > 0.0884), and those which were part of the liver fat CVAS to avoid sample overlap, resulting in 362,910 individuals.

To assess the relationship of rare inactivating variants with liver fat and related traits, we studied the subset of UK Biobank participants with exome sequencing data available. Sample quality control was performed by excluding samples that had no imputed genetic data, a genotyping call rate < 0.95, a mismatch between submitted and inferred sex, sex chromosome aneuploidy, exclusion from kinship inference, excessive third-degree relatives, or that were outliers in heterozygosity or genotype missingness rates, and restricting to samples of European ancestries as well as removing one of each pair of related individuals (2nd degree or closer, KING coefficient > 0.0884). We first analyzed the relationship between rare inactivating variants and liver fat in 11,021 individuals with both whole exome sequencing and abdominal MRI imaging data available. Next, to understand the relationship between inactivating variants in two genes, *APOB* and *MTTP*, and related biomarkers and disease states, we analyzed the full set of up to 42,260 participants with exome sequencing data available.

#### Framingham Heart Study

The Framingham Heart Study is a multigenerational prospective study that enrolled individuals free of cardiovascular disease beginning in 1948. 3,284 individuals in the Offspring and Third Generation cohorts (enrollment beginning in 1971 and 2002, respectively; Kannel et al., 1979; Splansky et al., 2007) with genotype data available underwent multidetector abdominal CT for liver fat quantification (Speliotes et al., 2008) and were included in analyses described below.

#### Multi-Ethnic Study of Atherosclerosis (MESA)

The Multi-Ethnic Study of Atherosclerosis (MESA) study is a prospective cohort that enrolled individuals free of cardiovascular disease between 2000 and 2002 (Bild et al., 2002). 4,195 individuals with genotype data available underwent multidetector CT for liver fat quantification (Zeb et al., 2012) and had genetic data available and were used in analyses described below.

#### Mass General Brigham Biobank

Mass General Brigham Biobank is a hospital-based biorepository with genetic data linked to clinical records (Karlson et al., 2016). Patients were defined as having NAFLD or NASH according to diagnosis codes in the electronic health care record (Supp. Table 2) and were compared to controls without such diagnoses as described below.

#### Informed Consent and Study Approval

The UK Biobank study was approved by the Research Ethics Committee (reference 16/NW/0274) and informed consent was obtained from all participants. Analysis of UK Biobank data was conducted under application 7089 and was approved by the Mass General Brigham institutional review board (protocol 2013P001840). Framingham Heart Study and MESA genotype and phenotype data were retrieved for analysis from NCBI dbGAP under procedures approved by the Mass General Brigham institutional review board (protocol 2016P002395). Mass General Brigham Biobank participants each provided written informed consent and analysis was approved by the Mass General Brigham institutional review board.

### Liver fat quantification in UK Biobank participants using a new machine learning algorithm

To determine liver fat percentage from abdominal magnetic resonance images, we used 2D Convolutional Neural Networks (CNNs) to estimate liver fat percentage from abdominal MRI in 38,706 individuals. The imaging protocol in UK Biobank was switched from gradient echo to IDEAL mid-study, and liver fat was previously quantified by Perspectum Diagnostics only in individuals imaged using the gradient echo protocol (Wilman et al., 2017). To be able to infer liver fat from both protocols, we therefore used a two-model approach with “teacher-student” models. The “teacher” model was a 2D CNN trained on 3,198 of the 4,412 individuals who underwent the gradient echo imaging protocol and had the full set of 10 standard images. The truth data for this model were liver fat values previously quantified by Perspectum Diagnostics from gradient echo imaging protocols which were made available to UK Biobank researchers. Validation was assessed in a held-out set of 1,214 participants with truth data. Liver fat values for the remaining 5,496 participants with gradient echo imaging were estimated using this model. To estimate liver fat in participants imaged using the IDEAL protocol, we also trained a “student” model in the participants who had undergone both the gradient echo and IDEAL imaging protocols. This model was also a 2D CNN, trained on 1,054 of the 1,437 overlapping samples. The truth data for this model was liver fat in the gradient echo protocol, which was inferred from the “teacher” model. The remaining 383 overlapping samples that were not used for training were used to validate the student model. Liver fat values for the remaining 28,595 participants with IDEAL imaging were inferred using this model. In total, we estimated liver fat for 34,281 participants with these models. Performance on the held-out test set is shown for each model in Supp. Figure 1.

The 2D CNNs were optimized with backpropagation and Adaptive Moment stochastic gradient descent (ADAM). The models were implemented in tensorflow version 2.1 using the ML4CVD modeling framework (Sarma et al., 2020). The python package hyperopt was used for Bayesian hyperparameter optimization of the model architecture to select the width, depth, activation function, and the size of each residual block in the CNN. The final architecture consisted of two layers of convolution followed by three residual blocks of 2 convolutions in parallel whose outputs are concatenated and max-pooled reducing the size of the representation by a factor of 4 after each block. The output of the final convolutional block is flattened and processed by two fully-connected layers and finally fed to the output regression neuron. All non-linear activations functions in the model are rectified linear units.

To combine the previously-quantified liver fat and results of the two models, we first used the previously-quantified liver fat estimates provided by the UK Biobank where available, including the n = 205 individuals who had a nonstandard number of gradient echo images and were not used in the ‘teacher’ model generation. When previously-quantified liver fat was unavailable, we preferentially used the liver fat estimates from the teacher model. When teacher model liver fat estimates were unavailable, we used the liver fat estimates from the student model. For subsequent analyses of liver fat, we filtered to 36,703 individuals in UK Biobank with genetic data, BMI measurements and liver imaging available. Final sources of liver fat were: n = 4,511 previously-quantified, n = 4,971 estimated from gradient echo protocol, n = 27,221 estimated from IDEAL protocol.

### Association of liver fat with patient baseline and clinical characteristics

Baseline characteristics of the 36,703 UK Biobank participants are shown in Supp. Table 1. Number of weekly drinks was defined in American standard drinks (1 drink = 14 g ethanol) according to the conversions: red or white wine, 0.84 drinks/glass; beer, 1.29 drinks/pint; liquor, 0.68 drinks/measure; fortified wine, 0.7 drinks/glass; other alcohol, 1 drink/glass. For participants who reported consuming alcohol monthly rather than weekly, monthly alcohol consumption was converted to weekly by multiplying by 0.23 (7×(365/12)). Excessive alcohol consumption was defined according to AASLD guidelines (Chalasani et al., 2012): greater than 14 weekly drinks if female or greater than 21 weekly drinks if male). Physician diagnosis of NAFLD and NAFLD/NASH were defined using ICD10 codes K76.0 or K76.0+K75.8, respectively. Other diseases were defined according to a combination of self-report, procedure codes and ICD codes (Supp. Table 2). Hepatic steatosis was defined as liver fat > 5.5%, as determined previously for UK Biobank using the original previously quantified liver fat values (Wilman et al., 2017). High waist-to-hip ratio was defined as greater than 0.9 if male and greater than 0.85 if female. Weight categories were defined using BMI: underweight, BMI< 18.5 kg/m^2^; normal, 18.5≤BMI< 25 kg/m^2^; overweight, 25≤BMI< 30 kg/m^2^; obese, 30≤BMI< 40 kg/m^2^; severely obese, BMI≥40 kg/m^2^.

To determine the clinical/anthropometric influences (sex, excessive alcohol consumption, physician diagnosis of NAFLD, physician diagnosis of diabetes) on median liver fat, or the effects of hepatic steatosis on median triglycerides, we performed median regression. Similarly, we used logistic regression to evaluate the effects of physician diagnosis of NAFLD on hepatic steatosis, and hepatic steatosis on diabetes or hypertension diagnosis. In both median and logistic regression, we included sex, birth year, age at imaging, age at imaging^2^ and MRI machine serial number as covariates. We calculated the Pearson correlation coefficient to assess the relationship between liver fat and BMI at time of imaging.

We also wanted to assess how imaging-based quantification of liver fat compared to predicting liver fat using clinical and anthropometric measurements. Liver fat percent is well-modeled by a beta distribution, as it is a series of proportions in the interval (0,1) (Ferrari and Cribari-Neto, 2004). We therefore constructed a beta regression model of liver fat using these measurements in the derivation and testing sets used to construct the ML model. We selected available anthropometrics, biomarkers associated with metabolic function and liver function or injury, as well as measurements of total body or abdominal fat available in UK Biobank. Lipid measures were adjusted for lipid-lowering medication use and blood pressure was adjusted for anti-hypertensive medication use, as previously described (Ehret et al., 2016; Patel et al., 2020). Missing values were imputed using *aregImpute* in the R package *Hmisc*. Only traits which were nominally (p< 0.05) associated with truth liver fat in univariable analysis were included in the beta regression model (Supp. Figure 2). Variables which were not associated with liver fat and were therefore excluded from the beta regression model were total bilirubin, direct bilirubin and indirect bilirubin. We constructed a variable beta regression model using 3,176 of the 4,412 individuals who underwent the gradient echo imaging protocol and had the full set of standard images. The truth data for this model were liver fat values previously quantified by Perspectum

Diagnostics. The variable beta regression model was constructed using the *betareg* package in R, optimizing the mean and precision link functions to logit and log, respectively, using AIC & BIC comparisons. Performance of the model was evaluated by the Pearson correlation between truth liver fat and predicted liver fat in a held-out testing dataset of 1,196 individuals who also had liver fat previously quantified by Perspectum Diagnostics (Supp. Figure 2).

### Genotyping and quality control

UK Biobank samples were genotyped on either the UK BiLEVE or UK Biobank Axiom arrays, then imputed into the Haplotype Reference Consortium and UK10K + 1000 Genomes panels. We excluded genotyped variants with call rate < 0.95, imputed variants with INFO score < 0.3, and imputed or genotyped variants with minor allele frequency less than 0.001 in the UK Biobank population. Variant positions were denoted in GRCh37/hg19 coordinates.

### Phenotype transformation

Because liver fat is not normally distributed and nor are its residuals with respect to clinical covariates, we transformed the input to a rank-based output. This approach is commonly used in GWAS of quantitative traits with skewed distributions (Yang et al., 2012). First, we took the residuals of liver fat in a linear model that included sex, year of birth, age at time of MRI, age at time of MRI squared, genotyping array, MRI device serial number, and the first ten principal components of ancestry. Then, we performed the inverse normal transform on the residuals from this model, yielding a standardized output with mean 0 and standard deviation of 1.

### Common variant association study

We performed a CVAS of the inverse normal transformed liver fat residuals in 32,976 individuals, applying linear mixed models with BOLT-LMM (version 2.3.4) to account for ancestry, cryptic population structure, and sample relatedness (Loh et al., 2015b). The default European linkage disequilibrium panel provided with BOLT was used. Variants with BOLT-LMM P < 5 × 10^−8^ were considered to be genome-wide significant. Loci were defined by 2 MB windows (1 MB distance from the most-significant variant in either direction). The most strongly associated variant at each locus is referred to as the lead variant. We determined the effects of each of the eight lead variants on liver fat % and presence of hepatic steatosis (liver fat > 5.5%) using linear and logistic regression, respectively, in the same 32,976 individuals in the CVAS, adjusting for sex, year of birth, age at time of MRI, age at time of MRI squared, genotyping array, MRI device serial number, and the first ten principal components of ancestry. We repeated this CVAS in the subset of 4,038 individuals with previously-quantified liver fat who passed the CVAS sample quality control.

We replicated the CVAS findings in the Framingham Heart Study and the Multi-Ethnic Study of Atherosclerosis (MESA). In the Framingham cohort (Offspring Cohort and Third Generation Cohort), we examined whether the 8 variants associate with hepatic steatosis on CT imaging. Genotyping was imputed to the HapRef consortium using the Michigan Imputation Server. After imputation, variants with allele frequency less than 0.01% and those with an imputation score of less than 0.3 were excluded from analysis. 3284 individuals in Framingham with genotype data available underwent multidetector abdominal CT (Speliotes et al., 2008). Hepatic steatosis was measured by computing the liver-to-phantom ratio of the average Hounsfield units of three liver measurements to average Hounsfield units of three phantom measurements (to correct for inter-individual differences in penetration), as previously described (Speliotes et al., 2008). We tested the association of all 8 variants with liver-to-phantom ratio with adjustment for age, sex and ten principal components of ancestry using a linear mixed model (BOLT-LMM) to control for relatedness among individuals. Liver fat measurements were inverse normal rank transformed prior to analysis.

In the Multi-Ethnic Study of Atherosclerosis cohort (MESA), genotypes were imputed to the HapRef consortium using the Michigan Imputation Server. After imputation, variants with allele frequency less than 0.01% and those with an imputation score of less than 0.3 were excluded from analysis. Hepatic steatosis was measured as the mean of three attenuation measurements, two in the right lobe of the liver and one in the left lobe (Zeb et al., 2012), without use of phantom measurement normalization. We therefore tested the association of the top CVAS variants with mean liver attenuation with adjustment for age, sex and ten principal components of ancestry. Mean liver attenuation was subject to inverse normal transformation prior to analysis. Liver fat measurements were inverse normal rank transformed prior to analysis. Linear regression with adjustment for age, sex and five principal components of ancestry was used to test for the association of genetic variants with liver fat. PLINK was used for analysis.

Individuals with higher liver fat have lower liver-to-phantom ratios and liver attenuation measurements. For interpretability, we therefore reported replication estimates in units of standard deviation increases in liver fat, with a one standard deviation increase in liver fat corresponding to a one standard deviation decrease in the liver-to-phantom ratio or mean liver attenuation.

We examined the association of the top CVAS variants with blood biomarkers alanine aminotransferase (ALT) and aspartate aminotransferase (AST) in UK Biobank using linear regression of each biomarker (in U/L) adjusting for sex, year of birth, age at enrollment and age at enrollment squared, genotyping array and the first ten principal components of ancestry.

We also examined the association of the top CVAS variants with physician diagnosis of NAFLD or NASH in UK Biobank and Mass General Brigham Biobank. Disease definitions are provided in Supp. Table 2. In UK Biobank, association of each top CVAS variant was assessed using logistic regression of disease status with sex, year of birth, age at enrollment and age at enrollment squared, genotyping array and the first ten principal components of ancestry as covariates. In the Mass General Brigham Biobank, genotyping was performed using an Illumina MEGA array. Variants were imputed to the HapRef consortium using the Michigan Imputation Server. Variants with multinucleotide alleles and those with call rate less than 90% were excluded prior to imputation. After imputation, variants with allele frequency less than 0.01% and those with an imputation score of less than 0.3 were excluded from analysis. Association of each top CVAS variant was assessed using logistic regression of disease status with age, sex and five principal components of ancestry as covariates.

### Polygenic score analysis

The 8 lead variants were scored in additive fashion based on number of liver-fat increasing variants identified weighted by their CVAS effect size estimates, producing a single score for each individual.

We tested for association between the score and incident disease occurrence after UK Biobank enrollment using a Cox model in the same set of individuals used to test associations between single CVAS variants and NAFLD/NASH. We excluded individuals who had any of the four diseases investigated, resulting in 362,096 participants in the analysis. We focused on the association of the score with liver diseases; given previously reported association of liver fat variants with circulating lipids (Liu et al., 2017), we also examined association of the score with circulating LDL cholesterol using linear regression. LDL cholesterol was adjusted for lipid-lowering medication use as previously described (Patel et al., 2020); liver disease definitions are listed in Supp. Table 2. All PRS analyses were adjusted for age at enrollment, sex, the first five principal components of ancestry, and genotyping array. We also quantified proportion of individuals who developed each disease during study follow-up stratified by PRS decile.

### Rare variant association study

In the subset of individuals with whole exome sequencing available, we identified rare (minor allele frequency < 0.001) inactivating variants in each gene. Sequencing data from the “Functionally Equivalent” gene sequencing dataset (https://biobank.ctsu.ox.ac.uk/showcase/label.cgi?id=170) was annotated using the Ensembl Variant Effect Predictor (VEP) software (version 96.0) LOFTEE. LOFTEE applies a set of filters to identify high-confidence inactivating variants based on predicted impact on the resulting transcript. High-confidence inactivating variants include those predicted to cause premature truncation of a protein (nonsense), insertions or deletions (indels) of DNA that scramble protein translation beyond the variant site (frameshift), or point mutations at sites of pre-messenger ribonucleic acid splicing that alter the splicing process (splice-site).

We aggregated the inactivating variants identified within each gene into a rare variant burden analysis: individuals were considered as an inactivating variant carrier for a particular gene if they had one or more loss-of-function mutations in the gene, and a non-carrier otherwise. We tested the association of inactivating variant carrier status for each gene with inverse normal transformed liver fat as previously described (see Phenotype Transformation) using linear regression with the first ten principal components of ancestry as covariates. We removed genes with fewer than 10 inactivating variant carriers to increase the likelihood of having sufficient statistical power to detect an effect, leaving 3,384 genes in the analysis. To determine the effects of *APOB* or *MTTP* inactivating variants on blood biomarkers or disease outcomes, we used linear or logistic regression, respectively, adjusting for sex, year of birth, age at enrollment and age at enrollment squared, genotyping array and the first ten principal components of ancestry. Lipid measures were adjusted for lipid-lowering medication use as previously described (Patel et al., 2020).

### Data availability

Summary statistics for the liver fat CVAS, as well as the machine learning model architectures and learned weights will be made available at the Cardiovascular Disease Knowledge Portal (http://broadcvdi.org/home/portalHome) and the ML4CVD modeling framework will be made available via GitHub repository at time of publication.

